# The effect of a mobile health application on enhancing adherence on ART among adolescents and young adults living with HIV: A Pilot Study in Fako Division, South West Region, Cameroon

**DOI:** 10.1101/2025.03.24.25324582

**Authors:** Charles Njumkeng, Tendongfor Nicholas, Prudence Tatiana Nti Mvilongo, Mbuwir Charlotte Bongfen, Elvis T Amin, Thomas Obinchemti Egbe, Patrick A. Njukeng

**Affiliations:** Global Health Systems Solutions, Douala, Cameroon; Department of Microbiology and Parasitology, University of Buea, Cameroon; Department of Public Health and Hygiene, University of Buea, Cameroon; Department of Gynaecology and Obstetrics, University of Buea

**Keywords:** Adherence, factors, efficiency, Antiretroviral Therapy, young adults, mHealth, adolescents, HIV, Cameroon

## Abstract

**Background:** Despite advancements in Antiretroviral Therapy (ART), adherence remains a challenge, particularly within younger populations. Considering that Adhering to lifelong ART treatment can often be challenging, this pilot study investigated the effect of mHealth application on adherence to ART among adolescents and young adults living with HIV in the Fako Division of Cameroon.

**Methods:** The study used a quasi-experimental design in which participants from selected treatment centers were proposed a mobile application to facilitate their adherence on treatment. Adherence measurements were taken before and after the intervention to assess the mobile application’s effectiveness. Adherence was measured using self-reports, pill counts, and viral load assessments.

**Results:** The study recruited 129 individuals, with more participants coming from Regional Hospital Limbe 49 (38.0%) and Regional Hospital Buea 47 (36.4%). Comparing adherence levels at baseline and end line, the most significant improvement was seen in pill count adherence, which rose from 56.8% to 79.1% (p<0.01). Increases in self-reported adherence and combined methods were also significant, with p-values of 0.003 and p<0.01, respectively. Viral suppression increased from 76.7% to 80.6%, but this change was not statistically significant (p=0.063).

**Conclusion:** The study’s findings indicate that the mHealth app effectively improves adherence to ART among adolescents and young adults, with significant increases observed after six months of use. This pilot study highlights the app’s potential to enhance care. However, further research with a larger sample size, longer duration, and a control group is recommended as next steps to assess the application’s impact on care.

## Background

The human immunodeficiency virus (HIV) remains one of the world’s most serious public health problems. It is estimated that about 39 million people are living with HIV globally, with 1.3 million people being newly infected with HIV and approximately 630,000 people died from AIDS-related illnesses in 2022[1,2]. The use of Antiretroviral Therapy (ART) has improved the quality of life for HIV patients across the globe. HIV-related morbidity and mortality have decreased, leading to improvements in the quality of life in countries where ART is widely available to People living with HIV.

High level of adherence to antiretroviral therapy among people living with HIV is critical in achieving optimal treatment outcomes. Poor adherence to ART can lead to treatment failure, viral resistance, higher opportunistic infections, increase disease transmission and increased morbidity and mortality[3–5].

With a focus on bringing the HIV pandemic under control by achieving viral suppression, countries including Cameroon took the commitment to launch the 95-95-95 initiative in 2020. The aspiration of this initiative is to achieve 95% viral suppression among people on ART[6]. Adherence to ART is directly associated with achieving viral suppression [7].

In Cameroon the gap of achieving these targets remains worrisome especially among the younger population. It is estimated that the number of HIV infection identified is 56.5% and 53.8% among adolescents aged <15 years and young adults aged 15-24 years, respectively, while viral suppression stands at 68% and 79% for the respective aged groups[8]. On the other hand the rate of retaining HIV diagnosed cases on treatment was 73.7% with the South West region being one of the regions with poor performance [9,10].

Achieving epidemic control will be more challenging if the challenges faced by younger people living with HIV, who will be on lifelong treatment, are not identified and provided with possible solutions. One of the major difficulties with this population is not testing to know their HIV status and their adherence to the HIV guidelines[11,12]. Considering that ART is a lifetime treating adherence to the life treatment is often a problem[3–5]. There is therefore need to continuously monitor adherence issues and its associated factors especially among the adolescents and young adults who will need to be on treatment for a longer period.

Mobile health (mHealth) has emerged as a transformative tool in enhancing healthcare delivery, particularly in resource-limited settings. By leveraging on mobile technology, healthcare providers can improve access to vital health information, facilitate timely communication, and streamline patient management [13–15]. This study aims to explore ways of developing and introducing mHealth application in the HIV program in an attempt to enhance adherence to treatment, monitor patient health, and provide real-time support and reminders for medication. By integrating mHealth solutions into HIV care, programs can improve healthcare outcomes, reduce stigma, and ultimately contribute to the achievement of the 95-95-95 targets.

## Methods Study Area

This study was conducted for 9 months from January 2024 to September 2024 in the Fako Division of the South West Region of Cameroon. Health facilities with the highest number of adolescents and young adults on HIV treatment in each of the four districts that make up the Fako Division were selected for the study. These include Muyuka District Hospital, Tiko District Hospital, Limbe Regional Hospital, and Buea Regional Hospital. Fako Division has many social and economic amenities and political institutions, which have contributed to its increasing population, the majority of whom were youths[16].

The South West Region has 36 HIV treatment centers located within 19 health districts, with 26,873 patients on ART. The prevalence of HIV in the region is estimated at 3.6% among individuals aged 15 years and above[17]. Data from the region showed that the positivity rate among adolescents aged 15-19 years is 3.3%, and 4.0% among young adults aged 20-24 years. Regarding the achievement of the UNAIDS 95-95-95 targets by 2030, it was estimated that 49,775 people were living with HIV in the region, of which approximately 91% had been identified, and only 67% of those identified had been initiated on treatment. On the other hand, viral suppression is estimated at 82% among people on ARV. In the South West Region, initiating adolescents and young adults on treatment and achieving viral suppression remain challenging.

### Study design

This study adopted a quasi-experimental design in which participants from selected treatment centers were given a mobile application to facilitate their adherence on treatment. Pre- and post-intervention measurements of adherence were conducted to assess the effectiveness of the mobile application.

#### 3.4 Target population

The population involved in this study consisted of adolescents and young adults on ART who were enrolled in one of the selected treatment centers in Fako Division. The study included participants aged 14 to 24 years of both sexes who had access to and/or owned a mobile phone and were willing to install the app before the start date of the study. We excluded participants who have been on ART for less than a year and those under 14 years old. Including participants who had used HAART for at least one year enabled us to calculate baseline indicators.

#### 3.7 Description of the Mobile health Application

The application was developed by a contracted team of Information Technology (IT) experts. The development was inspired by field challenges and insights gathered from Adolescents and young adults living with HIV and healthcare providers[18]. The application was designed with the aim to deliver personalized and tailored preventive and treatment services, including remote monitoring and follow-up for people living with HIV (PLHIV), in a safe, consistent, and timely manner.

##### 3.7.1 Functions of Mobile application

The application has three interfaces. The first interface contains general information about health and the app, as well as a login window. The second interface is the preventive arm, which provides information on HIV basics, myths versus facts, HIV transmission, HIV prevention, HIV stigma, and HIV testing. The treatment arm of the application allows for pill counting, daily pill alerts, interaction with healthcare providers regarding side effects and appointments, support groups, and the sharing of adherence tips. The preventive arm also allowed HIV-negative individuals to use the application. The use of the application by both HIV-positive and HIV-negative individuals prevented the perception that the app was exclusively for people living with HIV.

##### 3.7.2 Deployment and Monitoring of the intervention

The deployment began with comprehensive training sessions for personnel at the treatment centers, ensuring they were well-versed with the app’s functionalities and could assist clients in effectively using the app. The personnel were trained on the app’s installation, navigation of various options, and how to enroll participants. This included helping participants install the application and generate passwords for accessing each treatment center. Participants were then enrolled and trained in the usage of the application.

Continuous support and resources were provided to address any challenges that arose during implementation. A WhatsApp group was created for participants at each treatment center to facilitate communication and provide tailored guidance, helping to resolve their concerns and enhance user experience while fostering adherence to the intervention. Regular feedback on the application was collected from the WhatsApp group to monitor progress, allowing for real-time adjustments to improve effectiveness. The backend pill-tracking function was also monitored to ascertain whether the app was in use.

##### 3.7.3 Privacy and confidentiality

The app’s privacy and security framework is designed to provide robust protection for sensitive health data and ensure compliance with legal and ethical standards. Given that the app managed personal health data and enable communication between users and treatment centers, it was developed in compliance with various privacy and data protection laws. Some of the key privacy and confidentiality considerations for the app include: encryption, user authentication and authorization, secure login, data minimization and anonymization, and privacy by design and by default.

#### 3.8 Sample size determination

Sample size calculation was based on the null hypothesis that treatment outcomes at baseline and endpoint are equal. The primary measure of effect was viral suppression over six months. Based on information from the South west Regiona, it is estimated that approximately 69.5% of participants would achieve viral suppression without intervention, while the interventions would help 90% of the participants achieve viral suppression. A two-sided test was assumed, where an effect in either direction was interpreted at a significance level of 5%. The study used a power of 80% to yield a statistically significant result using a chi-squared test (assuming an intention-to-treat principle for the analysis) of the relative risk at alpha = 0.05 to detect a 21% difference (69% versus 90%). The formula for calculating the sample size was derived from an online sample size calculator [19]. The standard normal deviate for α = Zα = 1.9600

The standard normal deviate for β = Zβ = 0.8416 Pooled proportion = P = (q1*P1) + (q0*P0) = 0.4950

A = Zα√P(1-P)(1/q1 + 1/q0) = 1.9599

B = Zβ√P1(1-P1)(1/q1) + P0(1-P0)(1/q0) = 0.7749

C = (P1-P0)2 = 0.1521

Total group size = N = (A+B)2/C = 54

It was assumed that 10% of the participants could drop out of the study due to lost to follow-up or mortality. Therefore, a minimum of 60 participants was required for enrollment to allow for the 10% drop out.

#### 3.9 Sampling techniques

Within each of the four health districts that make up the Fako Division, one HIV testing and treatment center was selected for the study based on the enrollment of adolescents and young adults receiving treatment. Participants were chosen using a simple random sampling technique, with adolescents and young adults serving as the sampling frame. In each treatment center, participants’ names were generated. These names were then drawn by ballot to select individuals for the study. The number of participants from each treatment center was calculated as a proportion of the total number of adolescents and young adults receiving treatment at that center.

#### Adherence measurements

For the purpose of this study, adherence was measured using three different methods: self-report, pill count, and viral suppression. For self-report, medication adherence was assessed using the Morisky Medication Adherence Scale (MMAS), which is a generic self-reported medication-taking behavior scale. The MMAS consists of four items, each with a dichotomous answer (yes/no) [20]. For the pill count, participants were asked to bring their medication to the treatment center, where the number of pills remaining in the medication container was counted and matched with the expected remaining medication based on when they collected it. The number of consumed medications was calculated as a percentage, and any participants who had consumed at least 95% of the drugs were considered adherent. Lastly, participants’ viral load was measured, and those with fewer than 1,000 copies per milliliter were considered virally suppressed.

##### 3.12 Data Analysis

The data were transferred from Microsoft Excel 2013 to the Statistical Package for the Social Sciences (SPSS) version 25. The data were cleaned, and appropriate codes were applied to ensure accurate results during analysis. Descriptive variables such as participants’ age, health districts, means of transport, clinical factors, and more where summarized as percentages. McNemar’s Test was used to compare the adherence levels at baseline and at the end line with a significance level set at 0.05.

##### 3.13 Ethical considerations

Ethical clearance was obtained from the University of Buea Faculty of Health Sciences Institutional Review Board, with reference number 2023/187-01/UB/SG/IRB/FHS. Administrative authorization was granted by the Southwest Regional Delegation of Public Health and the District Health Services of the relevant districts. Written consent was obtained from adult participants, and written assent was obtained from the guardians/parents of minors, ensuring their understanding and voluntary participation in the study. The consent and assent forms were designed to include signed permission to review participants’ medical records during the study period. The protocol was registered under the Pan African Clinical Trial with the unique identification number for the registry: PACTR202310674176154.

## RESULTS

### 4.2.1 Sociodemographic characteristics of the study population

The study recruited 129 individuals, with more participants coming from Regional Hospital Limbe 49 (38.0%) and Regional Hospital Buea 47 (36.4%). Females constituted the majority of the study population, accounting for 77 (59.7%). Participants who had attained secondary education made up 98 (76.0%), while fewer participants had acquired at most primary education were the least 11(8.6%). In terms of occupation, the study was dominated by students 93 (72.1%), followed by employees 26 (20.3%). Over half (54.3%) of the participants were living with their biological parents, while those living with non-biological caregivers represented 33 (25.6%). With respect to age groups, the majority of the participants, 73 (56.6%), were adolescents aged 14 to 19 years, while 56 (43.4%) fell within the young adult age range of 20 to 24 years (Table 1). No loss to follow-up was recorded in the study.

**Table 1:** Sociodemographic characteristics of the study population.

| Variable | Characteristics | Frequency | Percentage |
| --- | --- | --- | --- |
| **Health District** | Tiko | 16 | 12.4 |
|  | Muyuka | 17 | 13.2 |
|  | Buea | 47 | 36.4 |
|  | Limbe | 49 | 38.0 |
|  | Total | 129 | 100.0 |
| **Sex** | Male | 52 | 40.3 |
|  | Female | 77 | 59.7 |
|  | Total | 129 | 100.0 |
| **Educational level** | Primary | 11 | 8.6 |
|  | Higher education | 20 | 15.5 |
|  | Secondary | 98 | 76.0 |
|  | Total | 129 | 100.0 |
| **Occupation** | Jobless | 10 | 7.8 |
|  | Employee | 26 | 20.3 |
|  | Student | 93 | 72.1 |
|  | Total | 129 | 100.0 |
| **Caregiver of adolescent** | Live alone | 26 | 20.1 |
|  | Non-biological parent | 33 | 25.6 |
|  | Biological parent | 70 | 54.3 |
|  | Total | 129 | 100.0 |
| **Age group** | ≤19years | 73 | 56.6 |
|  | 20 to 24years | 56 | 43.4 |
|  | Total | 129 | 100.0 |
| **Follow-up** | Loss to follow-up | 00 | 00.0 |
|  | Continued participation | 129 | 100.0 |
|  | Total | 129 | 100.0 |

### 4.2.2 Clinical factors of the study participants

From the clinical characteristics, it was observed that the majority of the participants initiated treatment before age 9, with 57 (44.2%) and 35 (27.1%) initiated treatment between the ages of 10 and 14. Fewer participants began treatment after age 20, with only 6 (4.7%). Regarding treatment duration, 48 participants (37.2%) had been on treatment for 5–9 years, while 47 (36.4%) had been on treatment for over 10 years, and 34 (26.4%) had been on treatment for less than 5 years. Most participants, 121 (93.8%), were taking first-line ART regimens. In terms of the age at which their status was disclosed, the majority were informed about their status between the ages of 10 and 14 years 79(61.2%), while a small number, 6 (4.7%), were disclosed at the time of diagnosis. Psychosocial support was made available to most participants, with 119 participants (92.2%) benefiting from it (Table 2).

**Table 2:** Clinical characteristics of the study population.

| **Variable** | **Characteristics** | **Frequency** | **Percentage** |
| --- | --- | --- | --- |
| Age at initiation | <9 | 57 | 44.2 |
|  | 10-14 | 35 | 27.1 |
|  | 15-19 | 31 | 24.0 |
|  | ≥20 | 6 | 4.7 |
|  | Total | 129 | 100.0 |
| Treatment duration (years) | < 5 years | 34 | 26.4 |
|  | 5 –9 years | 48 | 37.2 |
|  | >10 years | 47 | 36.4 |
|  | Total | 129 | 100.0 |
| ART regiment taking | First line | 121 | 93.8 |
|  | Second line | 8 | 6.2 |
|  | Total | 129 | 100.0 |
| Age at disclosure | 10-14 | 79 | 61.2 |
|  | 15-19 | 44 | 34.1 |
|  | At diagnosis | 6 | 4.7 |
|  | Total | 129 | 100.0 |
| psychosocial support | No | 10 | 7.8 |
|  | Yes | 119 | 92.2 |
|  | Total | 129 | 100.0 |
| Benefited workshop adherence on adherence | No | 51 | 39.5 |
|  | Yes | 78 | 60.5 |
|  | Total | 129 | 100 |

### 4.2.3 Measurement of Adherence rate

At baseline, adherence was measured using viral suppression, self-reporting, and pill count. Among the 129 participants enrolled in the study, 99 (76.7%) achieved viral suppression. The lowest adherence was recorded using the pill count method, with only 56.8% being adherent by this measure. It was observed that only 53 (41%) of participants were adherent across all three methods combined.

On the other hand at end line, the highest level of adherence was reported by self-reporting, with 106 (82.2%) of the 129 participants being adherent by this method. This was followed by viral suppression, with 104 (80.6%) of the participants achieving viral suppression. When considering adherence across all three methods (viral suppression, self-reporting, and pill count), only 81 (62.8%) of participants met the criteria for adherence (table 3).

**Table 3:** Measurement of Adherence rate at baseline.

| SN | Type of measurement | Baseline<br>N =129 (% , 95% CI) | End line<br>N =129 (% , 95% CI) | McNemar's<br>Test | P-value |
| --- | --- | --- | --- | --- | --- |
| 1 | Viral suppression (<1000 copies/ml) | 99(76.7; 69.4-84.0 ) | 104 (80.6; 73.8-87.5 ) | 3.21 | 0.063 |
| 2 | Adherence by self-reporting | 84(65.1; 56.9-73.3) | 106 (82.2; 75.6-88.8 ) | 8.820 | **0.003** |
| 3 | Adherence with Pill count (> 95%) | 73(56.8; 48.3-67.9) | 102 (79.1; 72.1-86.2 ) | 32.089 | **0.0001** |
| 4 | Adherence for all the three | 53 (41.00; 32.8-49.7 ) | 81 (62.8; 54.5-71.2 ) | 27.034 | **0.0001** |

### Comparison of adherence measurement at baseline and at end line

Comparing the levels of adherence at baseline and at end line, the highest change was observed in adherence measured by pill count, increasing from 56.8% at baseline to 79.1% at end line. This change was statistically significant (p<0.01). The increases in adherence levels measured by self-report and by all three methods combined were also significant, with p-values of 0.003 and 0.0001, respectively. With respect to viral suppression, there was an increase in viral suppression from 76.7% at baseline to 80.6% at end line. However, this increase was not significant (p = 0.063) (Table 3).

## Discussion

This study examined the effect of mHealth on enhancing adherence on ART among adolescent and young adults Living with HIV. At baseline, viral suppression was estimated to be 76.7%, which is very close to the national suppression rate for this age group, estimated at 76.2% [21]. In addition to the low viral suppression, adherence to ARTs, measured by pill count and self-reporting, was below 70%. This finding highlights the challenges the HIV program still faces in achieving the set objective of attaining epidemic control by 2030 within this age group, emphasizing the importance of seeking innovative means to improve service uptake among adolescents and young adults.

A study conducted in Yaoundé has shown a significant decrease in survival among adolescents on ART and suggested poor adherence to treatment as one of the main reasons [22]. Global adherence to ART among adolescents remains a challenge. Previous studies have reported suboptimal levels of adherence to treatment among adolescents in Cameroon[5,23–25]. The low adherence rate observed when the three methods were combined can indicate that, while some participants may be classified as adherent or as having viral suppression, the actual consistency of medication intake may be lower. The study’s findings showed an increase across all three methods used to measure adherence (viral suppression, self-reporting, and pill counts) after six months of using the mobile application. This suggests that the use of the mobile app had a positive effect on adherence among the study participants. Features of the application, such as reminders, educational resources, peer discussions, daily reminders and tracking, likely contributed in influencing participates behaviors leading to these improved outcomes.

This overall finding of this study is similar to previous studies conducted across different populations where mHealth applications have been examined in the context of enhancing HIV care. Mobile Health application was reported to positively influence linkage to treatment among Latino men who have sex with men[26], while in Nigeria, it was shown to increase the uptake of HIV self-testing services among young people[27]. In the U.S., the VIP-HANA app received promising feedback regarding its functions to assist people living with HIV in self-managing their symptom experiences by providing self-care strategies[28].

The significant improvements observed in the measurements of adherence by self-reporting and pill counts further suggest that the application has the ability to influence participant’s medications seeking behavior. It has been established that the measurement of adherence through self-reporting can provide information about personal or systemic barriers affecting adherence to treatment regimens, such as side effects, forgetfulness, or a lack of understanding of the importance of medication, as well as the patient’s motivation and engagement, support networks, or associated stigma [29,30]. These elements are directly targeted by our intervention through reporting effects, daily reminders, sharing adherence tips, and the social discussion function. Thus, they could possibly influence the observed changes in the measurement of adherence.

The observed increase in the proportion of viral suppression was not significant and can be explained by the fact that the study, at this stage, only explored the potential of the mobile app in supporting individuals in managing their health more effectively within six months of intervention, while the measurement of viral suppression may not capture short-term changes in adherence. A study conducted by the Centers for Disease Control and Prevention showed that their intervention resulted in a significant change in the level of viral suppression after a 12-month measurement period[31]. This further suggests that the six-month measurement of viral suppression might not have been sufficient to significantly change the proportion of viral suppression.

Previous works have reported that the level of adherence necessary for achieving viral suppression have showed considerable variation and presents different results based on the methods used to measure adherence [32]. Nonetheless, the observed no significant difference in the proportion of viral suppression in this study, compared to adherence measured by other methods, is similar to findings in other studies[32–34].

During the study period, we experienced no loss to follow-up. This suggests that the mobile health application may have enhanced engagement, keeping the participants connected throughout the study. Additionally, the app features were tailored to meet the needs of the participants, and regular communication with healthcare providers probably helped maintain their interest throughout the study. The regular reminders for medication intake, appointments, and WhatsApp group follow-ups may have contributed to reducing the likelihood of loss to follow-up. The discussion feature of the app also provided an avenue for peer connections, keeping the participants engaged. Participant engagement in the study can be supported by the overall significant increase in the level of adherence. This is in alignment with past studies that have shown that patients with poor or fair ART adherence were identified as being at risk of loss to follow-up [35].

## Conclusions

The findings of our study have shown that the mHealth app can improve overall adherence to ART among adolescents and young adults. Participants’ adherence measurements were significantly increased after six months of using the mobile application. This indicates the potential of the application in enhancing care, as demonstrated by this pilot study. However, we recommend further studies with a larger sample size, longer duration, and a control group to measure the impact of the application on enhancing care.

## Data Availability

The authors confirm that the data supporting the findings of the study are available within the article

## Acknowledgements

We express gratitude to the South West Regional Delegation of Public Health for facilitating the implementation of this study by providing administrative authorization. We salute the commitment of the various focal persons in the centers that were involved in the study. We also appreciate all participants for their collaboration in the implementation of the study.

## Authors’ contributions

CN conceived, designed and led the study implementation and data analysis, TN participated in the conception, designing and supervised the study implementation, PTNM participated in the designing and in developing the manuscript, MCB, participated in the designing and field implementation, ETA participated in the field implementation, TOE participated in the designing and supervised the study implementation PAN conceived, designed and supervised the study implementation. All authors read and approved the final manuscript.

## Disclaimer

All authors were involved in the study implementation.

